# HIV and viremia prevalence in non-migrating members of migrant households in Rakai region, Uganda: A cross-sectional population-based study

**DOI:** 10.1101/2023.12.08.23299745

**Authors:** Ruth Young, Joseph Ssekasanvu, Joseph Kagaayi, Robert Ssekubugu, Godfrey Kigozi, Steven J. Reynolds, Maria J. Wawer, Bareng A.S. Nonyane, Larry W. Chang, Caitlin E. Kennedy, Ligia Paina, Philip A. Anglewicz, Thomas C. Quinn, David Serwadda, Fred Nalugoda, M. Kate Grabowski

**Affiliations:** Department of International Health, The Johns Hopkins Bloomberg School of Public Health, Baltimore, MD, USA; Department of Epidemiology, The Johns Hopkins Bloomberg School of Public Health, Baltimore, MD, USA; Rakai Health Sciences Program, Kalisizo, Uganda; Makerere University School of Public Health, Kampala, Uganda; Division of Intramural Research, National Institute of Allergy and Infectious Diseases, National Institutes of Health, Bethesda, MD, United States; Department of Medicine, The Johns Hopkins School of Medicine, Baltimore, MD, USA; Department of Population, Family and Reproductive Health, The Johns Hopkins Bloomberg School of Public Health, Baltimore, MD, USA; Department of Pathology, The Johns Hopkins School of Medicine, Baltimore, MD, USA

**Keywords:** Transients and Migrants, Uganda, HIV, Family Relations, Prevalence

## Abstract

**Introduction:** In sub-Saharan Africa, migrants are more likely to be HIV seropositive and viremic than non-migrants. However, little is known about HIV prevalence and viremia in non-migrants living in households with in- or out-migration events. We compared HIV outcomes in non-migrating persons in households with and without migration events using data from the Rakai Community Cohort Study (RCCS), an open population-based cohort in Uganda

**Methods:** We analyzed RCCS survey data from one survey round collected between August 2016 and May 2018 from non-migrating participants aged 15–49. Migrant households were classified as those reporting ≥1 member moving into or out of the household since the prior survey. A validated rapid test algorithm determined HIV serostatus. HIV viremia was defined as >1,000 copies/mL. Modified Poisson regression was used to estimate associations between household migration and HIV outcomes, with results reported as adjusted prevalence ratios (adjPR) with 95% confidence intervals (95%CI). Analyses were stratified by gender, direction of migration (into/out of the household), and relationship between non-migrants and migrants (e.g., spouse).

**Results:** There were 14,599 non-migrants (7,654, 52% women) identified in 9,299 households. 4,415 (30%) lived in a household with ≥1 recent migrant; of these, 972(22%) had migrant spouses, 1,102(25%) migrant children, and 875(20%) migrant siblings. Overall, HIV prevalence and viremia did not differ between non-migrants in migrant and non-migrant households. However, in stratified analyses, non-migrant women with migrant spouses were significantly more likely to be HIV seropositive compared to non-migrant women with non-migrant spouses (adjPR:1.44, 95%CI:1.21-1.71). Conversely, non-migrant mothers living with HIV who had migrant children were less likely to be viremic (adjPR:0.34, 95%CI:0.13-0.86). Among non-migrant men living with HIV, spousal migration was associated with a non-significant increased risk of viremia (adjPR:1.37, 95%CI:0.94-1.99). Associations did not typically differ for migration into or out of the household.

**Conclusions:** Household migration was associated with HIV outcomes for certain non-migrants, suggesting that the context of household migration influences the observed association with HIV outcomes. In particular, non-migrating women with migrating spouses were more likely to have substantially higher HIV burden. Non-migrants with migrant spouses may benefit from additional support when accessing HIV services.

## Introduction

Across sub-Saharan Africa, migrants have higher HIV prevalence and incidence compared to non-migrants, and are less likely to be virally suppressed if living with HIV.[1–4] However, there is limited data on the impact of migration on HIV outcomes among non-migrants who remain behind or who welcome migrants into their households.

There is reason to believe that migration of a household member could lead to increased risk of HIV acquisition and viremia among non-migrants. Migration can be stressful due to changes in resources and relationships.[5–8] Studies have shown that some individuals cope with increased stress by adopting riskier sexual behaviors, which may result in higher risk of HIV acquisition.[9–12] Migration of a partner can precipitate or follow changes in relationship status.[5,13,14] Separation from partners[15,16], and reduced parental supervision[6] can also lead to risky sexual behaviors and, in turn, potential increased HIV acquisition risk. Furthermore, migration may change who in the household provides care to those living with HIV or what resources are available.[5] Caregivers are often family members provide a range of instrumental and emotional support, which has been linked to improved adherence to antiretroviral therapy.[17–21] Out-migration may result in changes to caregiving and may place those living with HIV at greater risk of viremia if there is reduced care coordination or less care support for the non-migrant.

The nature of the relationship between a migrant and a non-migrant may determine how disruptive migration is for the non-migrant, and, as a result, differentially influence HIV prevalence and viremia. Most prior studies examining the impact of migration on HIV among non-migrant residents have focused only on spouses with out-migrant partners.[15,16,22,23] Some studies found that non-migrant partners of migrants were more likely to report additional sexual partners[15,24] but this can vary by gender.[16] In addition, non-migrants with migrant partners were more likely to attribute additional HIV risk to their partner’s behavior rather than their own.[1,25] In Tanzania, Kishamawe et al. found higher HIV prevalence and incidence among men with wives who were mobile and frequently away compared to resident men with resident wives.[15] Few studies have considered how migration of other household members, such as children or siblings, may indirectly impact HIV outcomes.[18,26]

We evaluated whether non-migrants who experienced a recent migration event into or out of the household were more likely to be HIV seropositive and, if HIV seropositive, to be viremic, compared to residents in households with no recent history of migration. We stratified data by the relationship between the migrant and non-migrant to determine which relationships were more closely linked to HIV outcomes. We hypothesized that: (i) non-migrant women in households with a recent out-migration event were more likely to be HIV seropositive; and (ii) non-migrant men and women with migrant spouses would be more likely to be HIV seropositive and, if HIV seropositive, more likely to be viremic.

## Methods

### Study population and setting

We used cross-sectional data from the Rakai Community Cohort Study (RCCS) collected between August 2016 and May 2018 across 41 communities (37 inland, 4 fishing communities). The RCCS is an open population-based cohort set in a semi-rural region of south-central Uganda.[27] The RCCS first conducts a census to enumerate everyone in a household regardless of age and presence at time of census. The census collects information on their relationship to the head of the household and any movements into or out of the household since the last census. For migrants, a specific census mobility form records additional information including origin, destination, and reason for migration. Next, a survey collects demographic and self-reported sexual behaviors from residents between 15-49 years who have lived in the community for at least 6 months or at least one month with intention to stay longer. Duration of residency in the household and migration into or out of the household is confirmed at the time of survey. In addition, rapid HIV testing and counseling are offered, and blood samples are drawn to determine HIV status and viral load.

To evaluate the impact of migration on non-migrants, we restricted our analytic sample to non-migrating RCCS participants who consented to being surveyed and tested for HIV. Non-migrants were defined as survey-eligible participants who reported not moving or migrating since their last survey.

### Exposure

Our primary exposure of interest was migration in the household. In-migrants were defined as those who had migrated into the household from another community with the intention to stay or had already stayed for at least six months. Out-migrants were those who migrated out of the household to another community with the intention to stay in their destination community. Communities were classified based on community boundaries defined when the cohort was established in 1994. Migration was assessed using survey data for those eligible and using the mobility form in the census for everyone else. Individuals who reported moving within communities, circular migration, or returning to communities, were classified as mobile and excluded from the analysis. Such movements over smaller distances or return migration back to the community were theorized to be less disruptive than migrating between communities.

Households were categorized into four groups based on the direction of migration into the household: (i) no-migration households, where no recent migration event was documented; (ii) in-migration only households, where member(s) migrated into the household and no members migrated out; (iii) out-migration only households, where member(s) migrated out and no member(s) migrated into the household; and (iv) bidirectional migration households, where member(s) migrated in and member(s) migrated out.

### Relationship sub-analyses

Sub-analyses were conducted for different non-migrant and migrant relationships. Relationships were coded based on the most recent household roster (Table S1). Three relationships were evaluated: (i) non-migrants in long-term stable relationships (e.g., marriage), referred to as spouses, whose spouses migrate compared to non-migrants with spouses who do not migrate; (ii) non-migrant parents who have one or more migrant children compared to non-migrant parents whose children do not migrate; and (iii) non-migrants who have one or more migrant siblings compared to non-migrants whose siblings do not migrate. The direction of migration, i.e., into or out of the household, was not considered in relationship sub-analyses as the sample size was too small.

### Outcomes

Our primary outcomes were HIV seropositivity among all non-migrants and HIV viremia among HIV-positive non-migrants. HIV status was determined using a validated testing algorithm which included three rapid tests and enzyme immunoassays or polymerase chain reaction (PCR) confirmation.[28] HIV viral loads were measured using Abbott RealTime assays, and individuals with ≥1,000 copies/mL classified as having HIV viremia.[29]

### Statistical methods

We first compared the demographics and self-reported sexual behaviors across the four types of migrant households (no-migration, in-migration only, out-migration only and bidirectional) for men and women separately using Kruskal-Wallis tests and chi-square tests for continuous and categorical variables, respectively.

Next, we assessed the relationship between household migration and our two outcomes using univariate and multivariate Poisson regression models with robust standard errors.[30] Separate regressions were fitted for men and women because biological, social and structural correlates of HIV seropositivity and viremia differ between these groups.[31–34]. Measures of association were reported as prevalence ratios (PR) with 95% confidence intervals (95%CI).

To account for differences in demographics across no-, in-, out- and bi-directional households, we used multivariate regressions adjusted for demographic confounders based on observed differences across type of migrant household in bivariate analyses and previous research linking them to higher HIV prevalence. Potential confounders included age group[27], educational attainment, marital status[35], proportion of household members under 15 years, and residency in an inland or fishing community.[27]

We conducted several sensitivity analyses. For HIV viremia, we reduced the threshold from 1000 copies/mL to 400 copies/mL. To determine if the direction of migration in relationship sub-analyses mattered, we stratified relationship regressions by the direction of migration (in- or out-) and compared them to no-migration relationships. Relationship analyses were also stratified for fishing and inland communities, as fishing communities have substantially higher HIV prevalence and a different demographic profile than RCCS agrarian and trading communities.[27] Analyses used Stata 17[36] and R version 4.0.4.[37]

### Ethics

The RCCS protocol has approval from the Ugandan Virus Research Institute Research Ethics Committee (GC/127/19/11/137), Uganda National Council for Science and Technology (HS 540), Johns Hopkins School of Medicine IRB (IRB00217467), and Western IRB, Olympia Washington (Protocol #20031318). All participants provided written informed consent.

## Results

### Participant characteristics

At census, 22,086 non-migrant eligible residents out of 30,192 were identified, of whom 14,599 (7,654 women) participated in the RCCS and were included in the final analytic sample (Figure 1). Most individuals who did not participate were away for school or work (7,087/7,487; 95%). Of those present at time of survey, 243 (1.6%) refused to participate. Non-participants were significantly more likely to be men (58% vs. 48% p<0.001), living in fishing communities (23% vs. 22% p=0.019) and younger (median age: 25 vs. 31, p<0.001).

**Figure 1.**
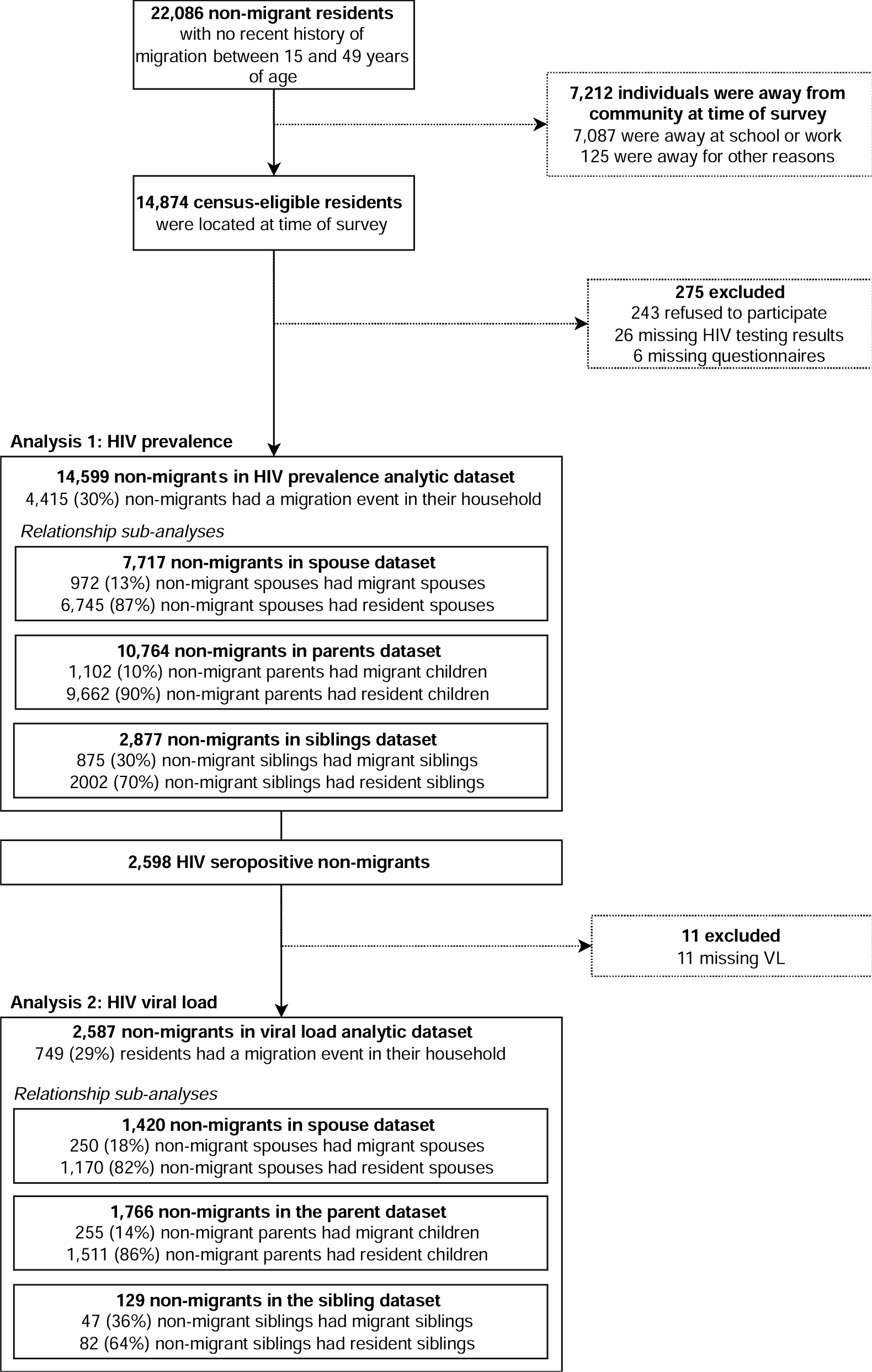
Study flowchart.

Of the 14,599 non-migrants, 70% (10,184) lived in households with no recent migration events (no-migration households). Among the remaining households, there were 4,064 migration events accounting for 21% of all migration events identified in the RCCS during the survey period (Figure S1, Figure 2). Most individuals lived in households with one (68.6%; 3030/4415) or two migration events (22.0%; 970/4415). Among the 30% (4,415/10,184) of participants living in households with at least one migration event, 9% (1,271) lived in in-migration households; 18% (2,558) in out-migration households; and 4% (586) were in bidirectional households.

**Figure 2.**
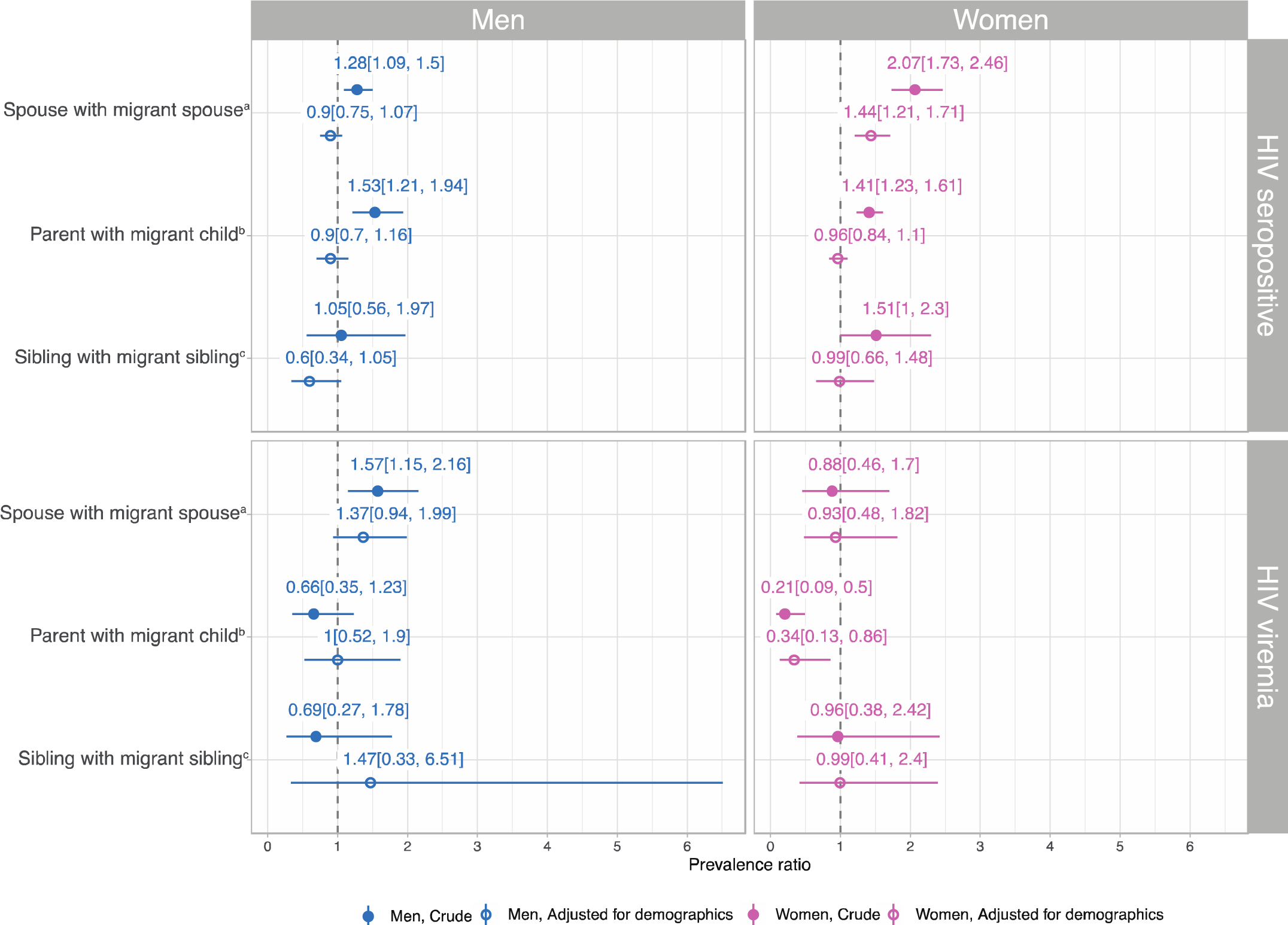
Crude and adjusted prevalence ratios for HIV status and HIV viremia by relationship between migrant and non-migrant. ^a^Compared to spouses with non-migrating spouses ^b^Compared to parents with non-migrating children ^c^Compared to siblings with non-migrating siblings Note: Prevalence ratios presented with 95% confidence intervals, parent and sibling regressions were adjusted for demographics (age, education, fishing community residence, proportion of household members under 15 years, marital status) and spouse regressions were adjusted for demographics except for marital status.

For both men and women, non-migrants in out-migration only households were younger and more likely to be previously married compared to non-migrants in no-migration households (Table 1). Sexual behaviors did not vary by type or directionality of migration within the household (Table S2).

**Table 1.** Demographic characteristics of 14,599 non-migrant residents in HIV prevalence analytic dataset stratified by presence and type of migration in the household.

|  | No migration<sup>a</sup> | In-migration only<sup>a</sup> | Out-migration only<sup>a</sup> | Bidirectional migration<sup>a</sup> |
| --- | --- | --- | --- | --- |
| **Men (n=6,945)** |  |  |  |  |
| Number of men | 4,722 | 693 | 1,202 | 328 |
| Education |  |  |  |  |
| None - primary | 3,263 (69.1) | 489 (70.6) | 824 (68.6) | 215 (65.5) |
| Secondary School | 1,172 (24.8) | 160 (23.1) | 324 (27.0) | 97 (29.6) |
| Beyond SS | 287 (6.1) | 44 (6.3) | 54 (4.5) | 16 (4.9) |
| Age (yrs) | 31 (23, 39) | 29 (23, 37) | 27 (18, 40) | 29 (22, 39) |
| Age category |  |  |  |  |
| 15-19 yrs | 807 (17.1) | 109 (15.7) | 345 (28.7) | 60 (18.3) |
| 20-24 yrs | 587 (12.4) | 112 (16.2) | 193 (16.1) | 47 (14.3) |
| 25-29 yrs | 674 (14.3) | 139 (20.1) | 116 (9.7) | 58 (17.7) |
| 30-34 yrs | 826 (17.5) | 115 (16.6) | 115 (9.6) | 52 (15.9) |
| 35-39 yrs | 748 (15.8) | 96 (13.9) | 116 (9.7) | 36 (11.0) |
| 40-44 yrs | 674 (14.3) | 72 (10.4) | 151 (12.6) | 41 (12.5) |
| 45-49 yrs | 406 (8.6) | 50 (7.2) | 166 (13.8) | 34 (10.4) |
| Marital status |  |  |  |  |
| Currently married | 2,626 (55.6) | 477 (68.8) | 421 (35.0) | 211 (64.3) |
| Prev. married | 615 (13.0) | 36 (5.2) | 228 (19.0) | 25 (7.6) |
| Never married | 1,481 (31.4) | 180 (26.0) | 553 (46.0) | 92 (28.0) |
| HH size | 4.0 (2.0, 6.5) | 3.5 (2.5, 6.5) | 5.5 (3.0, 8.0) | 5.0 (3.0, 8.5) |
| Proportion of HH <15 years | 0.4 (0.0, 0.6) | 0.4 (0.0, 0.5) | 0.3 (0.0, 0.5) | 0.3 (0.0, 0.5) |
| Resides in fishing community | 1,189 (25.2) | 237 (34.2) | 221 (18.4) | 107 (32.6) |
| **Women (n=7,654)** |  |  |  |  |
| Number of women | 5,462 | 578 | 1,356 | 258 |
| Education |  |  |  |  |
| None - primary | 3,378 (61.8) | 358 (61.9) | 853 (62.9) | 123 (47.7) |
| Secondary school | 1,714 (31.4) | 187 (32.4) | 435 (32.1) | 102 (39.5) |
| Beyond SS | 370 (6.8) | 33 (5.7) | 68 (5.0) | 33 (12.8) |
| Age (yrs) | 30 (23, 37) | 32 (24, 38) | 34 (20, 42) | 34 (24, 40) |
| Age category |  |  |  |  |
| 15-19 yrs | 796 (14.6) | 93 (16.1) | 317 (23.4) | 46 (17.8) |
| 20-24 yrs | 780 (14.3) | 61 (10.6) | 120 (8.8) | 21 (8.1) |
| 25-29 yrs | 1,005 (18.4) | 78 (13.5) | 92 (6.8) | 29 (11.2) |
| 30-34 yrs | 1,025 (18.8) | 111 (19.2) | 159 (11.7) | 38 (14.7) |
| 35-39 yrs | 897 (16.4) | 118 (20.4) | 213 (15.7) | 55 (21.3) |
| 40-44 yrs | 645 (11.8) | 75 (13.0) | 258 (19.0) | 41 (15.9) |
| 45-49 yrs | 314 (5.7) | 42 (7.3) | 197 (14.5) | 28 (10.9) |
| Marital status |  |  |  |  |
| Currently married | 3,450 (63.2) | 341 (59.0) | 576 (42.5) | 133 (51.6) |
| Prev. married | 936 (17.1) | 111 (19.2) | 328 (24.2) | 59 (22.9) |
| Never married | 1,076 (19.7) | 126 (21.8) | 452 (33.3) | 66 (25.6) |
| HH size | 5.0 (3.0, 7.0) | 5.5 (3.5, 7.0) | 6.5 (4.5, 8.0) | 7.0 (5.0, 9.5) |
| Proportion of HH <15 years | 0.5 (0.3, 0.6) | 0.4 (0.3, 0.6) | 0.4 (0.3, 0.5) | 0.4 (0.3, 0.5) |
| Resides in fishing community | 1,029 (18.8) | 157 (27.2) | 162 (11.9) | 42 (16.3) |

SS = Secondary School; yrs = years; Prev. = previously; HH= household
<sup>a</sup>Statistics presented: n(%); Median (Q1, Q3)

### Prevalence of HIV among non-migrants in households with migration and without migration

The overall prevalence of HIV among non-migrants was 18% (2,598/14,599), with women more likely than men to be living with HIV (20% vs. 15%; PR:1.29 (95%CI:1.20-1.39)). Depending on the direction of migration, non-migrant women in-migration households had a prevalence of HIV ranging from 15-25% compared to 20% in non-migrant women in no-migration households. For non-migrant men living in households with migration, HIV prevalence ranged from 15-17% compared to 16% in no-migration households (Table 2).

**Table 2.** Crude and adjusted prevalence ratios for HIV status and HIV viremia among non-migrating persons aged 15-49 in households which experienced a migration event.

|  | HIV seropositive |  |  | HIV viremia |  |  |
| --- | --- | --- | --- | --- | --- | --- |
|  | n/N(%) | Crude PR<br>(95%CI) | Adjusted PR<br>(95%CI) | n/N(%) | Crude PR<br>(95%CI) | Adjusted PR<br>(95%CI) |
| **Men** |  |  |  |  |  |  |
| No-migration (ref) | 752/4722<br>(16) | 1<br>[1,1] | 1<br>[1,1] | 139/1089<br>(13) | 1<br>[1,1] | 1<br>[1,1] |
| In-migration only | 120/693<br>(17) | 1.09<br>[0.91,1.30] | 1.04<br>[0.89,1.22] | 11/140<br>(7.9) | 0.93<br>[0.65,1.33] | 0.94<br>[0.663,1.325] |
| Out-migration only | 150/1202<br>(12) | **0.78\*\***<br>**[0.67,0.92]** | 0.96<br>[0.83,1.12] | 21/255<br>(8.2) | 0.95<br>[0.69,1.30] | 0.87<br>[0.64,1.19] |
| Bidirectional | 49/328<br>(15) | 0.94<br>[0.72,1.22] | 0.93<br>[0.73,1.18] | 4/39<br>(10) | 1.20<br>[0.76,1.90] | 1.3<br>[0.79,2.13] |
| Observations | 6945 | 6945 | 6945 | 1064 | 1064 | 1064 |
| P-value<sup>a</sup> | 0.006 |  |  | 0.10 |  |  |
| **Women** |  |  |  |  |  |  |
| No-migration (ref) | 1090/5462<br>(20) | 1<br>[1,1] | 1<br>[1,1] | 182/749<br>(24) | 1<br>[1,1] | 1<br>[1,1] |
| In-migration only | 142/578<br>(25) | **1.23\*\***<br>**[1.06,1.43]** | 1.05<br>[0.92,1.21] | 27/119<br>(23) | 0.62<br>[0.34,1.11] | 0.73<br>[0.41,1.31] |
| Out-migration only | 256/1356<br>(19) | 0.95<br>[0.84,1.07] | 0.93<br>[0.83,1.04] | 34/148<br>(23) | 0.65<br>[0.42,1.00] | 0.7<br>[0.45,1.08] |
| Bidirectional | 39/258<br>(15) | 0.76<br>[0.56,1.02] | 0.78<br>[0.59,1.02] | 14/48<br>(29) | 0.80<br>[0.31,2.06] | 0.82<br>[0.33,2.04] |
| Observations | 7654 | 7654 | 7654 | 1523 | 1523 | 1523 |
| P-value<sup>a</sup> | 0.012 |  |  | 0.80 |  |  |

Exponentiated coefficients, prevalence ratios; 95% confidence intervals in brackets
\* p < 0.05, \*\* p < 0.01, \*\*\* p < 0.001
<sup>a</sup>Statistical tests performed: chi-square test of independence
<sup>b</sup>4 men and 7 women were missing HIV viral loads measures.
Model: adjusted for demographics (age category, education status, fishing or inland community, proportion of household members under 15 years, marital status)

### Prevalence of HIV among non-migrants by relationship to migrating household member

Next, we assessed whether HIV prevalence varied by relationship between the non-migrant and migrant. In most cases, crude HIV seroprevalence was higher among non-migrants with migrant spouses and children, and non-migrant women with migrant siblings (Table S3). We identified 972(19%) non-migrants with migrant spouses, 1,102(22%) with migrant children, and with 875(17%) migrant siblings. Most other non-migrant participants had either non-familial relationships (970, 19%) or relationships that could not be classified (Table S1).

We found that HIV seroprevalence was significantly higher among non-migrant women with migrant spouses compared to married non-migrant women without migrant spouses (36% vs. 17%; adjPR:1.44; 95%CI:1.21-1.71, p<0.001; Figure 2, Table S3). This positive association between HIV seroprevalence and spousal migration was strong among non-migrant women living in inland (adjPR:1.79; 95%CI:1.37-2.33) whereas in fishing communities this association was no longer observed (adjPR:1.20; 95%CI:0.97-1.47, Table S5). Non-migrant men and women with migrant spouses often cited relationship formation or dissolution as the reason for migration (79% (768/972)). However, when restricting analyses to currently married non-migrant women only, women with migrant spouses were still more likely to be HIV seropositive (adjPR:1.25; 95%CI:0.98-1.59, p=0.07, Table S5). In analyses stratified by direction of migration, spouse outmigration was more strongly associated with HIV seroprevalence compared to spouse in-migration among non-migrant women (Figure 3).

**Figure 3.**
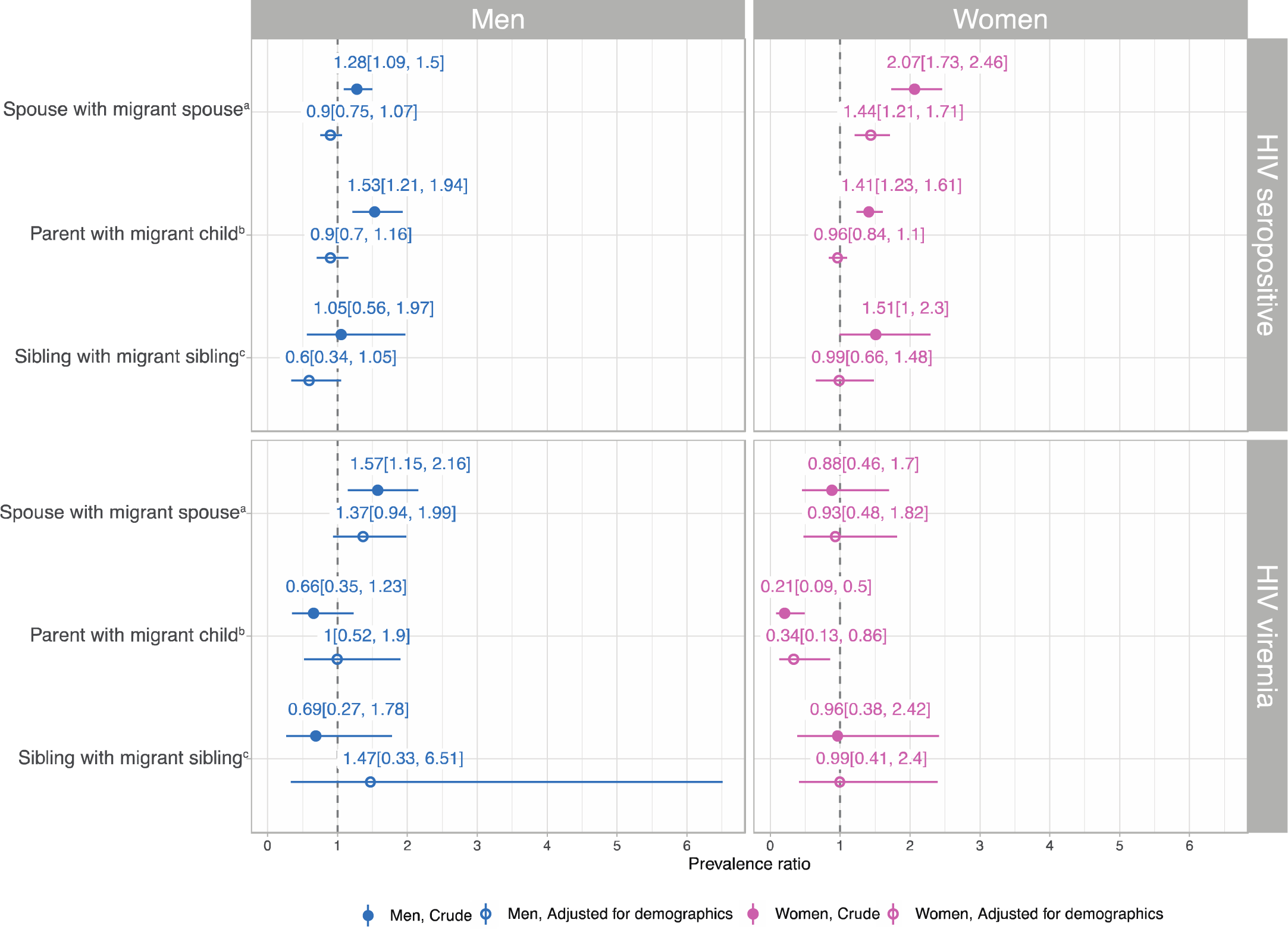
Crude and adjusted prevalence ratios for HIV and HIV viremia for relationship sub-analyses stratified by the direction of migration. ^a^Compared to spouses with non-migrating spouses ^b^Compared to parents with non-migrating children ^c^Compared to siblings with non-migrating siblings Note: Prevalence ratios presented with 95% confidence intervals, parent and sibling regressions were adjusted for demographics (age, education, fishing community residence, proportion of household members under 15 years, marital status) and spouse regressions were adjusted for demographics except for marital status. Sibling regressions for HIV viremia were excluded due to limited sample size.

Non-migrant men with migrant spouses also had higher HIV seroprevalence than non-migrant men with spouses who did not migrate; however, after adjustment for non-migrant participant demographics there was no longer any association between spousal migration and HIV serostatus. Similarly, child and sibling migration was not associated with HIV serostatus among non-migrant men or women after adjustment for non-migrant participant demographics (Figure 2). Parent and sibling regressions were adjusted for age, education, fishing community residence, proportion of household members under 15 years, marital status, and spouse regressions were adjusted for demographics except for marital status.

### Prevalence of HIV viremia among non-migrants in households with migration and without migration

For viremia, 16.7% (432/2587) of HIV-seropositive non-migrants were viremic, with men more likely to be viremic than women (PR:2.10 95%CI:1.76-2.51). In adjusted analyses, direction of migration in the household was not associated with HIV prevalence irrespective of gender (Table 2). Among non-migrant men and women living with HIV, prevalence of viremia did not vary across direction of migration in the household in either crude or adjusted analyses.

### Prevalence of HIV viremia among non-migrants by relationship to migrating household member

Prevalence of HIV viremia among HIV-positive non-migrants also varied by the relationship between non-migrant and migrants (Figure 2). For example, non-migrant men with migrant spouses were more likely to be viremic compared to non-migrant men with non-migrant spouses (28% vs. 18%; PR:1.57; 95%CI:1.15-2.16; adjPR:1.37; 95%CI:0.94-1.99; Table S3), and this did not vary by direction of spousal migration (Figure 3, Table S6). In contrast, non-migrant mothers with migrant children were significantly less likely to be viremic (3% vs. 13%; adjPR:0.34; 95%CI:0.13-0.86, Table S3), although few (n=5/135) viremic non-migrant mothers had a migrant child.

## Discussion

We found that the relationship between household migration and non-migrant’s HIV outcome depends on the context (e.g., relationship between migrant and non-migrant, and gender). In most cases, there was no relationship. However, spouse and child migration were linked to HIV prevalence and viremia: non-migrant women with migrant spouses were more likely to be living with HIV, and non-migrant men with migrant spouses were somewhat more likely to be viremic. In contrast, non-migrant mothers with migrant children were significantly less likely to be viremic. These findings suggest that the relationship between the migrant and the non-migrant is key to understanding the impact of household migration on HIV outcomes.

Our results indicate that spouse migration is associated with higher prevalence of HIV for non-migrant women. As our analysis was cross-sectional, it is unclear if HIV seroconversion predates spouse migration so spouse migration may lead to higher risk of HIV acquisition, or if those living with HIV have a higher risk of spouse migration. Previous studies in Africa have suggested that non-migrant women with migrant spouses are more likely to be HIV seropositive as they and their partners are more likely to engage in sexual behavior that predisposes them to greater risk of acquiring HIV.[15,22] Similarly, as spouse migration is often associated with relationship formation and dissolution, our results may reflect changes in relationships. Marital dissolution is associated with higher HIV incidence[35] and greater instability for women living with HIV, leading to spouse migration.[13,38]

Our findings also suggest that non-migrant men with migrant spouses may be more likely to be viremic regardless of whether spouses migrated in or out of the household. Different demographics could explain the higher rates of HIV viremia, as non-migrant men with migrant spouses tended to be younger and less educated. Alternatively, non-migrant men with migrant spouses may be less likely to engage in HIV treatment, if spouse migration is associated with relationship breakdown, or if a wife migrated for work, challenging traditional masculine roles and norms.[39] Men who feel that their masculinity is challenged by spouse migration may compensate by outwardly asserting masculine characteristics and avoid health clinics or taking medication, or experience more internalized stigma.[39,40] Alternatively, men with migrant spouses may be more stressed and have less time[41,42] to access health services or obtain antiretroviral therapy. Additional qualitative research is needed to better understand what men experience preceding, during, and following spouse migration.

We found an inverse relationship between child migration and HIV viremia among non-migrant women, suggesting that migration of a child may facilitate engagement in HIV care. Based on existing studies that highlight how children support parents living with HIV[43,44], we suggest several potential reasons why migrant children may reduce prevalence of viremia among their parents. Out-migrating children could increase the wealth of the household through remittances or improve intrahousehold resource allocation as the number of dependents decrease.[43–45] In-migrating children may provide additional assistance in the household, act as care providers for mothers or supportive care for other dependents in the household.[17,43,46] As this phenomena was not observed among fathers with migrant children, it may reflect gendered roles within the household. Compared to men, women take on more informal care within a household,[47] and so in- or out-migrating children may influence the mother’s time and resources, directly impacting access to HIV care. Alternatively, households with migrant children may be wealthier or more educated in ways not captured by our analysis and so individuals in those households may be virally suppressed regardless of whether children migrate. If older children are increasingly providing care support for their parents living with HIV, interventions designed to support caregivers may be needed. Further context-specific research would be needed to understand how child migration changes household dynamics, especially caregiving roles, and how that impacts access to healthcare.

Furthermore, our findings suggest additional characteristics to consider when examining how migration is linked with HIV outcomes for non-migrants. First, non-migrant and migrant relationships associated with changes in relationship status or sexual networks are associated with prevalence of HIV. For viremia, changes to relationships more likely to be linked to changes in caregiving, such as non-migrant parents with migrant children, may facilitate or impede access to treatment.[18,26] Second, the direction of migration alone was less important than we had hypothesized. Third community characteristics will impact the observed relationship between migration and HIV outcomes. In communities where HIV is common, like fishing communities in this region, the difference in HIV prevalence between non-migrant and migrant households may be less. Similarly, in communities where spouse migration is common, such as rural Mozambique, the association between spouse migration and HIV prevalence may be weaker.[48]

Our work has several limitations. The analysis excluded migration events within communities or circular migration (e.g., due to work or polygamous relationships). Separate examination of circular migration of spouses is warranted as it implies regular and returning movement between several places, distinct from other types of migration which are unidirectional. Previous research has also found an association between circular migration and HIV risk among left-behind spouses, as circular migrants are likely to travel home frequently, bridging sexual networks, and are more likely to have concurrent partners.[49,50]. Second, the data did not include potential confounders like individual wealth and changes in support (e.g. monetary, social support) provided to the non-migrant. Third, our data suggests that the impact of migration on non-migrants is likely context-specific and our findings may not apply to other contexts which have different kinds of migration and HIV epidemics. Fourth, to classify the relationship between migrant and non-migrant, we relied on the reported relationship to the head of household, but this may not capture all relationships within the household. Similarly, reported relationships may not truly reflect the support (e.g. caregiving, financial) provided.[51] We assumed that familial relationships within the household provided caregiving and support, but sources outside the household may also provide these.[52,53] Fifth, few parent-child relationships were identified in our data likely due to age-eligibility for the cohort and school-related absence during data collection. Previous literature has suggested that parental migration may impact sexual behaviors and HIV status of children remaining behind.[54,55] Lastly, analyses were prespecified, we cannot rule out that these findings were due to chance.

HIV prevention or treatment interventions have rarely been designed to incorporate migrant family members, although supportive relationships can facilitate HIV prevention and treatment.[56–58] Our results suggest that, for HIV outcomes, interventions may benefit from considering specific migrant relationships. For example, interventions tailored for couples with one migrant spouse may improve outcomes for both non-migrants and migrants. Identifying relationships where one partner is a migrant may be difficult; community health workers who may be more familiar with households could be best positioned to identify such couples. Counselling for those with migrant spouses could focus on how couple dynamics change, coping with stress, and planning for mutual support; this may support treatment adherence or reduce behaviors associated with HIV acquisition.[59,60]

Further, interventions targeting couples with one migrant partner may need to consider the specific needs of non-migrating men. Qualitative research on how household relationships and masculine identities change for non-migrant men with migrant spouses could inform future interventions by providing support and messaging that connect HIV care with masculine ideals. For instance, compared to women, men are less likely to join peer support groups as they perceive it to contradict masculine ideals of respectability, strength, and financial responsibility.[33,61,62] By incorporating income-generating activities or other opportunities to support or transform masculine ideals in peer support groups, men may be more likely to participate and improve treatment adherence.[62] In addition, a range of differentiated service delivery models could improve adherence by increasing access to services through extended opening hours or additional access points for working men.[63]

### Conclusions

Household migration is associated with HIV outcomes for certain sub-populations, particularly non-migrants with migrant spouses. Our findings suggest that when designing interventions to improve HIV outcomes, non-migrants with migrant spouses may benefit from targeted support when accessing HIV services.

## Declarations

### Ethics approval and consent to participate

The Rakai Community Cohort Study protocol has approval from the Ugandan Virus Research Institute Research Ethics Committee (GC/127/19/11/137), Uganda National Council for Science and Technology (HS 540), Johns Hopkins School of Medicine IRB (IRB00217467), and Western IRB, Olympia Washington (Protocol #20031318). All participants provided written informed consent. All methods were carried out in accordance with relevant guidelines and regulations. The secondary data analysis presented in this article was determined to not be human subjects research as it used deidentified, existing RCCS data by the Johns Hopkins Bloomberg School of Public Health Institutional Review Board.

### Consent for publication

Not applicable.

### Availability of data and materials

A de-identifed version of the data can be provided to interested parties subject to completion of the Rakai Health Sciences Program data request form and signing of a Data Transfer Agreement. Inquiries should be directed to.

## Authors’ contributions

RY and MKG conceived and designed the study. RY, MKG, JS, JK, RS, GK, SJR, MJW, BAS, LWC, CEK, LP, PAA, TCQ, DS, and FN contributed to the acquisition, analysis, and interpretation of the data for the study. RY wrote the initial draft manuscript. RY, JS, JK, RS, GK, SJR, MJW, BAS, LWC, CEK, LP, PAA, TCQ, DS, FN, and MKG contributed to the draft of the manuscript, provided substantial inputs, critical comments and suggested additional analyses. RY and MKG finalized the manuscript. All authors read and approved the final manuscript.

## Acknowledgements

We thank the study participants and community leaders in the Rakai Community Cohort Study, the Rakai Health Sciences Program staff, and our funders without whom this research would not be possible.

## Authors’ information

This work was presented at the Conference on Retroviruses and Opportunistic Infections (CROI), online, February 12 to 16, 22, 23, and 24, 2022.

## List of abbreviations

95%CI: 95% confidence interval
adjPR: Adjusted prevalence ratio
ART: Antiretroviral therapy
HH: Household
HIV: Human immunodeficiency virus
mL: Milliliter
PCR: Polymerase chain reaction
PR: Crude prevalence ratio
RCCS: Rakai Community Cohort Study
SS: Secondary school
yrs: years

## References

[1] Dzomba A, Tomita A, Govender K, Tanser F. Effects of Migration on Risky Sexual Behavior and HIV Acquisition in South Africa: A Systematic Review and Meta-analysis, 2000-2017. AIDS and Behavior 2018;1:1–35. 10.1007/s10461-018-2367-z.

[2] UN General Assembly. Political Declaration on HIV and AIDS: On the Fast Track to Accelerating the Fight against HIV and to Ending the AIDS Epidemic by 2030. Resolution 70/266, United Nations; 2016.

[3] Olawore O, Tobian AAR, Kagaayi J, Bazaale JM, Nantume B, Kigozi G, et al. Migration and risk of HIV acquisition in Rakai, Uganda: a population-based cohort study. The Lancet HIV 2018;5:e181–9. 10.1016/S2352-3018(18)30009-2.

[4] Billioux VG, Chang LW, Reynolds SJ, Nakigozi G, Ssekasanvu J, Grabowski MK, et al. Human immunodeficiency virus care cascade among sub-populations in Rakai, Uganda: an observational study 2017. 10.7448/IAS.20.1.21590.

[5] Lu Y. Household migration, social support, and psychosocial health: The perspective from migrant-sending areas. Social Science & Medicine 2012;74:135–42.

[6] Wu LL, Martinson BC. Family Structure and the Risk of a Premarital Birth. American Sociological Review 1993;58:210–32.

[7] Cavanagh SE, Fomby P. Family Instability in the Lives of American Children. Annual Review of Sociology 2019;12:41. 10.1146/annurev-soc-073018.

[8] Fomby P, Cherlin AJ. Family Instability and Child Well-Being. Am Sociol Rev 2007;72:181–204.

[9] Hulland EN, Brown JL, Swartzendruber AL, Sales JM, Rose ES, Diclemente RJ. The association between stress, coping, and sexual risk behaviors over 24 months among African-American female adolescents. Psychology, Health and Medicine 2015;20:443–56. 10.1080/13548506.2014.951369.

[10] Turpin R, Brotman RM, Miller RS, Klebanoff MA, He X, Slopen N. Perceived stress and incident sexually transmitted infections in a prospective cohort. Annals of Epidemiology 2019;32:20–7. 10.1016/j.annepidem.2019.01.010.

[11] Nansel TR, Riggs MA, Yu KF, Andrews WW, Schwebke JR, Klebanoff MA. The association of psychosocial stress and bacterial vaginosis in a longitudinal cohort. American Journal of Obstetrics and Gynecology 2006;194:381–6. 10.1016/j.ajog.2005.07.047.

[12] Amabebe E, Anumba DOC. Psychosocial stress, cortisol levels, and maintenance of vaginal health. Frontiers in Endocrinology 2018;9. 10.3389/fendo.2018.00568.

[13] Porter L, Hao L, Bishai D, Serwadda D, Wawer MJ, Lutalo T, et al. HIV status and union dissolution in Sub-saharan Africa: The case of Rakai, Uganda. Demography 2004;41:465–82. 10.1353/dem.2004.0025.

[14] Cassels S, Jenness SM, Khanna AS. Conceptual Framework and Research Methods for Migration and HIV Transmission Dynamics. AIDS Behav 2014;18:2302–13. 10.1007/s10461-013-0665-z.

[15] Kishamawe C, Vissers DCJ, Urassa M, Isingo R, Mwaluko G, Borsboom GJJM, et al. Mobility and HIV in Tanzanian couples: Both mobile persons and their partners show increased risk. Aids 2006;20:601–8. 10.1097/01.aids.0000210615.83330.b2.

[16] Palk L, Blower S. Mapping divided households and residency changes: the effect of couple separation on sexual behavior and risk of HIV infection. Scientific Reports 2015;5:17598–17598. 10.1038/srep17598.

[17] Angotti N, Mojola SA, Schatz E, Williams JR, Gómez-Olivé FX. ‘Taking care’ in the age of AIDS: older rural South Africans’ strategies for surviving the HIV epidemic. Culture, Health & Sexuality 2018;20:262–75. 10.1080/13691058.2017.1340670.

[18] Skovdal M. Examining the trajectories of children providing care for adults in rural Kenya: Implications for service delivery. Children and Youth Services Review 2011;33:1262–9. 10.1016/j.childyouth.2011.02.023.

[19] Ssekubugu R, Renju J, Zaba B, Seeley J, Bukenya D, Ddaaki W, et al. “He was no longer listening to me”: A qualitative study in six Sub-Saharan African countries exploring next-of-kin perspectives on caring following the death of a relative from AIDS. AIDS Care 2019;31:754–60. 10.1080/09540121.2018.1537467.

[20] Knodel J, Kespichayawattana J, Saengtienchai C, Wiwatwanich S. The role of parents and family members in ART treatment adherence: Evidence from Thailand. Res Aging 2010;32:19–39. 10.1177/0164027509348130.

[21] Damulira C, Mukasa MN, Byansi W, Nabunya P, Kivumbi A, Namatovu P, et al. Examining the relationship of social support and family cohesion on ART adherence among HIV-positive adolescents in southern Uganda: baseline findings. Vulnerable Child Youth Stud 2019;14:181–90. 10.1080/17450128.2019.1576960.

[22] Lurie MN, Williams BG, Khangelani Z, Mkaya-Mwamburi D, Garnett GP, Sweat MD, et al. Who infects whom? HIV-1 concordance and discordance among migrant and non-migrant couples in South Africa. AIDS 2003;17:2245–52. 10.1097/00002030-200310170-00013.

[23] Agadjanian V, Arnaldo C, Cau B. Health Costs of Wealth Gains: Labor Migration and Perceptions of HIV/AIDS Risks in Mozambique. Social Forces 2011;89:1097–117.

[24] Lurie MN, Williams BG, Zuma K, Mkaya-mwamburi D, Garnett GP, Sturm AW, et al. The Impact of Migration on HIV-1 Transmission in South Africa. Sexually Transmitted Diseases 2003;30:149–56. 10.1097/00007435-200302000-00011.

[25] Dladla AN, Hiner CA, Qwana E, Lurie M. Speaking to rural women: The sexual partnerships of rural South African women whose partners are migrants. Society in Transition 2001;32:79–82. 10.1080/21528586.2001.10419032.

[26] Lowenthal ED, Marukutira T, Tshume O, Chapman J, Nachega JB, Anabwani G, et al. Parental Absence From Clinic Predicts Human Immunodeficiency Virus Treatment Failure in Adolescents. JAMA Pediatrics 2015;169:498–500. 10.1001/jamapediatrics.2014.3785.

[27] Chang LW, Quinn TC, Reynolds SJ, Health Sciences Program R, W Chang UL, Grabowski MK, et al. Heterogeneity of the HIV epidemic in agrarian, trading, and fishing communities in Rakai, Uganda: an observational epidemiological study. Lancet HIV 2016;3. 10.1016/S2352-3018(16)30034-0.

[28] Kagulire SC, Opendi P, Stamper PD, Nakavuma JL, Mills LA, Makumbi F, et al. Field evaluation of five rapid diagnostic tests for screening of HIV-1 infections in rural Rakai, Uganda. Int J STD AIDS 2011;22:308–9. 10.1258/ijsa.2009.009352.

[29] WHO | Consolidated guidelines on the use of antiretroviral drugs for treating and preventing HIV infection: what’s new n.d.

[30] Zou G. A Modified Poisson Regression Approach to Prospective Studies with Binary Data. Am J Epidemiol 2004;159:702–6. 10.1093/aje/kwh090.

[31] Scully EP. Sex Differences in HIV Infection. Curr HIV/AIDS Rep 2018;15:136–46. 10.1007/s11904-018-0383-2.

[32] Sia D, Onadja Y, Hajizadeh M, Heymann SJ, Brewer TF, Nandi A. What explains gender inequalities in HIV/AIDS prevalence in sub-Saharan Africa? Evidence from the demographic and health surveys. BMC Public Health 2016;16:1136. 10.1186/s12889-016-3783-5.

[33] Sileo KM, Fielding-Miller R, Dworkin SL, Fleming PJ. A scoping review on the role of masculine norms in men’s engagement in the HIV care continuum in sub-Saharan Africa. AIDS Care 2019;31:1435– 46. 10.1080/09540121.2019.1595509.

[34] Gupta GR, Parkhurst JO, Ogden JA, Aggleton P, Mahal A. Structural approaches to HIV prevention. The Lancet 2008;372:764–75. 10.1016/S0140-6736(08)60887-9.

[35] Nalugoda F, Guwatudde D, Bwaninka JB, Makumbi FE, Lutalo T, Kagaayi J, et al. Marriage and the Risk of Incident HIV infection in Rakai, Uganda. J Acquir Immune Defic Syndr 2014;65:91–8. 10.1097/QAI.0b013e3182a7f08a.

[36] StataCorp. Stata Statistical Software: Release 17 2021.

[37] R Core Team. R: A language and environment for statistical computing. 2021.

[38] Anglewicz P, Reniers G. HIV Status, Gender, and Marriage Dynamics among Adults in Rural Malawi. Studies in Family Planning 2014;45:415–28. 10.1111/j.1728-4465.2014.00005.x.

[39] Sileo KM, Reed E, Kizito W, Wagman JA, Stockman JK, Wanyenze RK, et al. Masculinity and engagement in HIV care among male fisherfolk on HIV treatment in Uganda. Culture, Health and Sexuality 2018;1058. 10.1080/13691058.2018.1516299.

[40] Sileo KM, Wanyenze RK, Mukasa B, Musoke W, Kiene SM. The Intersection of Inequitable Gender Norm Endorsement and HIV Stigma: Implications for HIV Care Engagement for Men in Ugandan Fishing Communities. AIDS Behav 2021. 10.1007/s10461-021-03176-1.

[41] Deng R, Lyttleton C. Linked spaces of vulnerability: HIV risk amongst migrant Dai women and their left-behind husbands in Southwest China. Culture, Health Sexuality 2013;15. 10.1080/13691058.2013.772241.

[42] Yeoh BSA, Somaiah BC, Lam T, Acedera KF. Doing Family in “Times of Migration”: Care Temporalities and Gender Politics in Southeast Asia. Ann Am Assoc Geogr 2020;110:1709–25. 10.1080/24694452.2020.1723397.

[43] Nalugya R, Russell S, Zalwango F, Seeley J. The role of children in their HIV-positive parents’ management of antiretroviral therapy in Uganda. African Journal of AIDS Research 2018;17:37–46. 10.2989/16085906.2017.1394332.

[44] Abimanyi-Ochom J, Inder B, Hollingsworth B, Lorgelly P. Invisible work: Child work in households with a person living with HIV/AIDS in Central Uganda. SAHARA-J: Journal of Social Aspects of HIV/AIDS 2017;14:93–109.

[45] Kohler IV, Kohler H-P, Anglewicz P, Behrman JR. Intergenerational transfers in the era of HIV/AIDS: Evidence from rural Malawi. Demogr Res 2012;27:775–834.

[46] Schatz E, David I, Angotti N, Gómez-Olivé FX, Mojola SA. From “Secret” to “Sensitive Issue”: Shifting Ideas About HIV Disclosure Among Middle-Aged and Older Rural South Africans in the Era of Antiretroviral Treatment. J Aging Health 2021:08982643211020202. 10.1177/08982643211020202.

[47] Schatz E, Seeley J. Gender, ageing & carework in East and Southern Africa: A review. Glob Public Health 2015;10:1185–200. 10.1080/17441692.2015.1035664.

[48] Agadjanian V, Hayford SR. Men’s Migration, Women’s Autonomy, and Union Dissolution in Rural Mozambique. Journal of Family Issues 2018;39:1236–57. 10.1177/0192513X17698184.

[49] Lurie M, Harrison A, Wilkinson D, Karim SA. Circular migration and sexual networking in rural KwaZulu/Natal: implications for the spread of HIV and other sexually transmitted diseases. Health Transition Review 1997;7:17–27.

[50] Martins-Fonteyn E, Loquiha O, Baltazar C, Thapa S, Boothe M, Raimundo I, et al. Factors influencing risky sexual behaviour among Mozambican miners: a socio-epidemiological contribution for HIV prevention framework in Mozambique. International Journal for Equity in Health 2017;16:179– 179. 10.1186/s12939-017-0674-z.

[51] Manderson L, Block E. Relatedness and care in Southern Africa and beyond. Social Dynamics 2016;42:205–17. 10.1080/02533952.2016.1218139.

[52] Madhavan S, Clark S, Beguy D, Kabiru CW, Gross M. Moving beyond the household: Innovations in data collection on kinship. Population Studies 2017;71:117–32. 10.1080/00324728.2016.1262965.

[53] Madhavan S, Clark S, Araos M, Beguy D. Distance or location? How the geographic distribution of kin networks shapes support given to single mothers in urban Kenya. Geographical Journal 2018;184:75–88. 10.1111/geoj.12230.

[54] Odimegwu CO, Wet ND, Adedini SA, Appunni S. Family Demography in Sub-Saharan Africa: Systematic Review of Family Research. In: Odimegwu CO, editor. Family Demography and Post-2015 Development Agenda in Africa, Cham: Springer International Publishing; 2020, p. 9–56. 10.1007/978-3-030-14887-4_2.

[55] Albert LM, Edwards J, Pence B, Speizer IS, Hillis S, Kahn K, et al. Associations of Father and Adult Male Presence with First Pregnancy and HIV Infection: Longitudinal Evidence from Adolescent Girls and Young Women in Rural South Africa (HPTN 068). AIDS Behav 2021;25:2177–94. 10.1007/s10461-020-03147-y.

[56] Mukumbang FC, Knight L, Masquillier C, Delport A, Sematlane N, Dube LT, et al. Household-focused interventions to enhance the treatment and management of HIV in low- and middle-income countries: a scoping review. BMC Public Health 2019;19:1–14. 10.1186/s12889-019-8020-6.

[57] Lauffenburger JC, Khan NF, Brill G, Choudhry NK. Quantifying Social Reinforcement Among Family Members on Adherence to Medications for Chronic Conditions: a US-Based Retrospective Cohort Study. Journal of General Internal Medicine 2019;34:855–61. 10.1007/s11606-018-4654-9.

[58] Masquillier C, Wouters E, Mortelmans D, Wyk B van, Hausler H, Damme WV. HIV/AIDS Competent Households: Interaction between a Health-Enabling Environment and Community-Based Treatment Adherence Support for People Living with HIV/AIDS in South Africa. PLOS ONE 2016;11:e0151379. 10.1371/journal.pone.0151379.

[59] Ruark A, Kajubi P, Ruteikara S, Green EC, Hearst N. Couple Relationship Functioning as a Source or Mitigator of HIV Risk: Associations Between Relationship Quality and Sexual Risk Behavior in Peri-urban Uganda. AIDS Behav 2018;22:1273–87. 10.1007/s10461-017-1937-9.

[60] Conroy AA, McKenna SA, Comfort ML, Darbes LA, Tan JY, Mkandawire J. Marital infidelity, food insecurity, and couple instability: A web of challenges for dyadic coordination around antiretroviral therapy. Social Science and Medicine 2018;214:110–7. 10.1016/j.socscimed.2018.08.006.

[61] Chikovore J, Gillespie N, McGrath N, Orne-Gliemann J, Zuma T. Men, masculinity, and engagement with treatment as prevention in KwaZulu-Natal, South Africa. AIDS Care 2016;28:74–82. 10.1080/09540121.2016.1178953.

[62] Mburu G, Ram M, Siu G, Bitira D, Skovdal M, Holland P. Intersectionality of HIV stigma and masculinity in eastern Uganda: implications for involving men in HIV programmes. BMC Public Health 2014;14:1–9. 10.1186/1471-2458-14-1061.

[63] Sharma M, Barnabas RV, Celum C. Community-based strategies to strengthen men’s engagement in the HIV care cascade in sub-Saharan Africa. PLOS Medicine 2017;14:e1002262. 10.1371/journal.pmed.1002262.

