## Additional file for "HIV and viremia prevalence in non-migrating members of migrant households in Rakai region, Uganda: A cross-sectional population-based study"

Additional file includes:

**Table S1** Coding for relationship between migrant and non-migrant.
**Figure S1** Sankey diagram of potential types of migration and inclusion/exclusion in analysis.
**Table S2** Sexual behaviors and sexually transmitted infections for 14,599 non-migrants in HIV prevalence analytic dataset stratified for men and women and by type of migration in the household.
**Table S3** Crude and adjusted prevalence ratios for HIV prevalence and HIV viremia with household migration stratified by the relationship between migrant and non-migrant.
**Table S4** Prevalence of HIV and viremia among spouses restricted to currently married only.
**Table S5** Crude and adjusted prevalence ratios for HIV and viremia regressions for relationship between migrant and non-migrant stratified by fishing and inland community.
**Table S6** Crude and adjusted prevalence ratios for HIV and HIV viremia by relationship between migrant and non-migrant stratified by direction of migration (into or out).

**Table S1 Coding for relationship between migrant and non-migrant.**

| | | Non-migrant and migrant relationships based on HH roster | | Number of unique non-migrants |
|---|---|---|---|---|
| Relationship code | Migrant | n | % | n |
| 1 | Child is the migrant, parent is the non-migrant | 1,407 | 21.87% | 1,102 |
| 2 | Parent moves, child is the non-migrant | 127 | 1.97% | 113 |
| 3 | Partner/spouse migrates, partner/spouse is the non-migrant | 1111 | 17.27% | 972 |
| 4 | Sibling moves, sibling non-migrant | 1083 | 16.83% | 875 |
| 5 | Grandchild moves, grandparent non-migrant | 20 | 0.31% | 17 |
| 6 | Grandparent moves, grandchild non-migrant | 3 | 0.05% | 3 |
| 8 | Migrant is related but not captured in the categories above e.g., uncle or aunt | 1,308 | 20.33% | 1,043 |
| 9 | Migrant is unrelated to non-migrant | 1,020 | 15.85% | 783 |
| 99 | It is unclear if the migrant and non-migrant are related | 355 | 5.52% | 187 |
| | Total | 6,434 | | 5,095 |

*Coding*

To determine the relationship between migrant and non-migrant, census household rosters listing the relationship of each member to the head of household were used in conjunction with spousal relationships from the questionnaire. We collapsed some relationship categories (e.g. biological child, stepchild) assuming some relationships had the similar roles. We reclassified relationships as follows:

- Co-wives were reclassified as spouses to the head of household;
- Parents and parents-in-law as parents;
- Biological son or daughter, son or daughter-in-law, and adopted, foster or stepchild as children.

The table above summarizes the nine different relationships coded between migrant and non-migrant. Relationships were classified based on the current household roster where the relationship was listed. If that was not possible, then the prior household roster was used to classify the relationship between the migrant and non-migrant. All relationships that were classified based on the previous round were outmigrant relationships

*Summary of relationships between migrant and non-migrants*

In total there were 6,434 unique migrant and non-migrant pairs. Parent migration may be more common than presented in the table above as those under 15 years were not eligible for the survey.

**Figure S1 Sankey diagram of potential types of migration and inclusion/exclusion in analysis.**

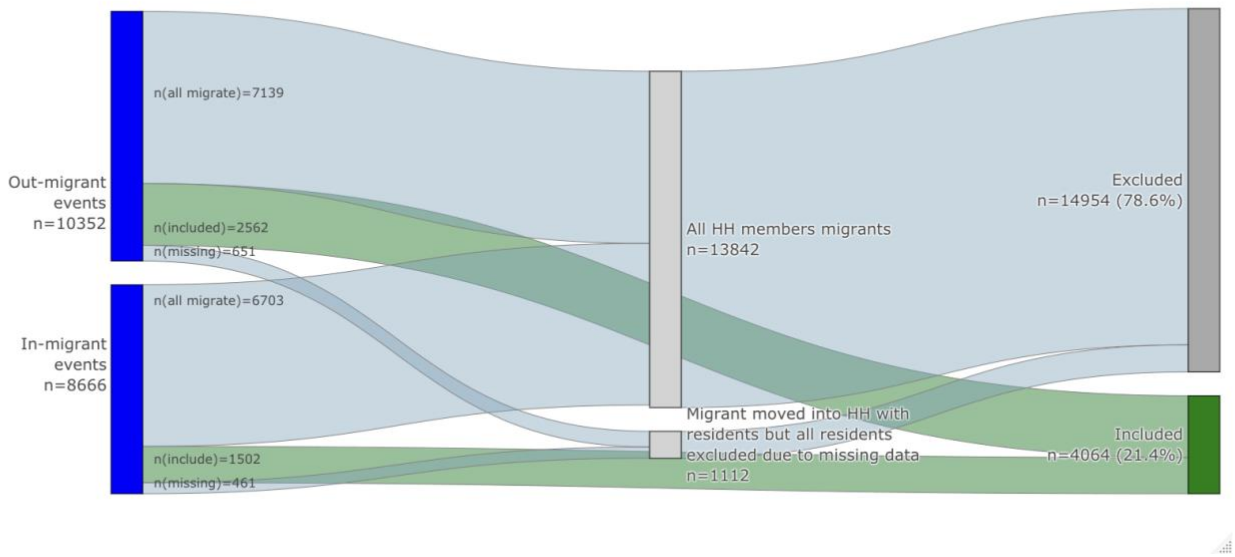


Note: HH - Household

19,018 migration events were recorded in the survey and census. Of these 73% (13,842/19,018) were migrants who moved into or out of households where all members were migrants and so were excluded from the study. An additional 5.85% (1,112/19,018) of migration events were excluded as the migrant moved into a home with census-eligible RCCS residents but the residents in those household did not participate in the RCCS. Overall, 21.4% (4,064/19,018) of migration events were captured in our analysis.

**Table S2 Sexual behaviors and sexually transmitted infection symptoms for 14,599 non-migrants in HIV prevalence analytic dataset stratified for men and women and by type of migration in the household.**

| | No migration[a] | In-migration only[a] | Out-migration only[a] | Bidirectional migration[a] | p-value[b] |
|---|---|---|---|---|---|
| Women (n=7,654) | | | | | |
| *Number of sexual partners in past year* | | | | | 0.047 |
| 0-1 partners | 5,094/5,462 (93) | 535/578 (93) | 1,288/1,356 (95) | 236/258 (91) | |
| 2+ partners | 368/5,462 (6.7) | 43/578 (7.4) | 68/1,356 (5.0) | 22/258 (8.5) | |
| *Genital ulcer in past year* | 485/5,451 (8.9) | 49/578 (8.5) | 85/1,352 (6.3) | 12/258 (4.7) | 0.002 |
| (Missing) | 11 | 0 | 4 | 0 | |
| *Sexually active* | 4,421/5,462 (81) | 454/578 (79) | 905/1,356 (67) | 183/258 (71) | <0.001 |
| *Casual partners, condom use*[c] | | | | | <0.001 |
| Stable partners only | 3,210/4,421 (73) | 321/454 (71) | 551/905 (61) | 124/183 (68) | |
| Casual partners, consistent condom use | 267/4,421 (6.0) | 25/454 (5.5) | 91/905 (10) | 13/183 (7.1) | |
| Casual partners, inconsistent condom use | 944/4,421 (21) | 108/454 (24) | 263/905 (29) | 46/183 (25) | |
| Men (n=6,945) | | | | | |
| *Number of sexual partners in past year* | | | | | <0.001 |
| 0-1 partners | 2,839/4,722 (60) | 430/693 (62) | 794/1,202 (66) | 178/328 (54) | |
| 2+ partners | 1,883/4,722 (40) | 263/693 (38) | 408/1,202 (34) | 150/328 (46) | |
| *Genital ulcer symptoms in past year* | 140/4,722 (3.0) | 21/693 (3.0) | 32/1,202 (2.7) | 19/328 (5.8) | 0.028 |
| (Missing) | | | | | |
| *Sexually active* | 3,828/4,722 (81) | 577/693 (83) | 833/1,202 (69) | 269/328 (82) | <0.001 |
| *Casual partners, condom use*[c] | | | | | <0.001 |
| Stable partners only | 1,644/3,828 (43) | 301/577 (52) | 302/833 (36) | 124/269 (46) | |
| Casual partners, consistent condom use | 661/3,828 (17) | 79/577 (14) | 165/833 (20) | 42/269 (16) | |
| Casual partners, inconsistent condom use | 1,523/3,828 (40) | 197/577 (34) | 366/833 (44) | 103/269 (38) | |
| *Circumcised man* | 2,992/4,722 (63) | 469/693 (68) | 793/1,202 (66) | 226/328 (69) | 0.022 |

Note:
[a] Statistics presented: n/N (%)
[b] Statistical tests performed: chi-square test of independence
[c] Amongst sexually active (had sex in the past year) participants only.
[d] Amongst HIV seropositive individuals only.

**Table S3 Crude and adjusted prevalence ratios for HIV prevalence and HIV viremia with household migration stratified by the relationship between migrant and non-migrant.**

| | Exposed n/N | Reference n/N | Crude PR [95%CI] | p | Adjusted PR [95%CI] | p |
|---|---|---|---|---|---|---|
| **HIV prevalence** | | | | | | |
| **Men** | | | | | | |
| Spouse with migrant spouse[a] | 158/706 | 482/2754 | 1.28 [1.09, 1.50] | <0.01 | 0.90 [0.75, 1.07] | 0.22 |
| Parent with migrant child[b] | 65/346 | 487/3972 | 1.53 [1.21, 1.94] | <0.01 | 0.90 [0.70, 1.16] | 0.40 |
| Sibling with migrant sibling[c] | 14/490 | 29/1067 | 1.05 [0.56, 1.97] | 0.88 | 0.60 [0.34, 1.05] | 0.07 |
| **Women** | | | | | | |
| Spouse with migrant spouse[a] | 95/266 | 690/3991 | 2.07 [1.73, 2.46] | <0.01 | 1.44 [1.21, 1.71] | <0.01 |
| Parent with migrant child[b] | 193/756 | 1030/5690 | 1.41 [1.23, 1.61] | <0.01 | 0.96 [0.84, 1.10] | 0.60 |
| Sibling with migrant sibling[c] | 33/385 | 53/935 | 1.51 [1.00, 2.30] | 0.05 | 0.99 [0.66, 1.48] | 0.95 |
| **HIV viremia** | | | | | | |
| **Men** | | | | | | |
| Spouse with migrant spouse[a] | 44/156 | 86/480 | 1.57 [1.15, 2.16] | <0.01 | 1.37 [0.94, 1.99] | 0.10 |
| Parent with migrant child[b] | 9/63 | 105/484 | 0.66 [0.35, 1.23] | 0.19 | 1.00 [0.52, 1.90] | 1.00 |
| Sibling with migrant sibling[c] | 4/14 | 12/29 | 0.69 [0.27, 1.78] | 0.44 | 1.47 [0.33, 6.51] | 0.61 |
| **Women** | | | | | | |
| Spouse with migrant spouse[a] | 9/94 | 75/690 | 0.88 [0.46, 1.70] | 0.71 | 0.93 [0.48, 1.82] | 0.83 |
| Parent with migrant child[b] | 5/192 | 130/1027 | 0.21 [0.09, 0.50] | <0.01 | 0.34 [0.13, 0.86] | 0.02 |
| Sibling with migrant sibling[c] | 6/33 | 10/53 | 0.96 [0.38, 2.42] | 0.94 | 0.99 [0.41, 2.40] | 0.99 |

[a]Compared to spouses with non-migrating spouses
[b]Compared to parents with non-migrating children
[c]Compared to siblings with non-migrating siblings
Note: Prevalence ratios (PR) presented with 95% confidence intervals (CI), spouse regressions adjusted for demographics except for marital status; parent and sibling regressions adjusted for demographics including marital status.

**Table S4 Prevalence of HIV and viremia among spouses restricted to currently married only.**

| | | HIV seropositive | | | | HIV viremia >1000 copies/mL | | | HIV viremia >400 copies/mL | | | |
|---|---|---|---|---|---|---|---|---|---|---|---|---|
| | | PR | 95% CI | p-value | N | PR | 95% CI | p-value | PR | 95% CI | p-value | N |
| **Men** | | | | | | | | | | | | |
| Crude | All spouses[a] | 1.28 | [1.09 ,1.50] | 0.003 | 3460 | 1.57 | [1.15 ,2.16] | 0.005 | 1.15 | [1.12 ,2.07] | 0.008 | 636 |
| Crude | Currently married only[a] | 1.25 | [1.05 ,1.50] | 0.014 | 3286 | 1.46 | [1.01 ,2.09] | 0.042 | 1.01 | [1.00 ,2.02] | 0.051 | 595 |
| Adjusted | All spouses[a] | 0.90 | [0.75 ,1.07] | 0.223 | 3460 | 1.37 | [0.94 ,1.99] | 0.104 | 0.94 | [0.93 ,1.92] | 0.118 | 636 |
| Adjusted | Currently married only[a] | 0.91 | [0.75 ,1.09] | 0.307 | 3286 | 1.27 | [0.85 ,1.89] | 0.252 | 0.85 | [0.84 ,1.82] | 0.281 | 595 |
| **Women** | | | | | | | | | | | | |
| Crude | All spouses[a] | 2.07 | [1.73 ,2.46] | <0.01 | 4257 | 0.88 | [0.46 ,1.70] | 0.705 | 0.97 | [0.54 ,1.76] | 0.927 | 784 |
| Crude | Currently married only[a] | 1.86 | [1.46 ,2.36] | <0.01 | 4138 | 0.76 | [0.29 ,2.00] | 0.583 | 0.69 | [0.26 ,1.80] | 0.448 | 735 |
| Adjusted | All spouses[a] | 1.44 | [1.21 ,1.71] | <0.01 | 4257 | 0.93 | [0.48 ,1.82] | 0.833 | 1.02 | [0.56 ,1.88] | 0.946 | 784 |
| Adjusted | Currently married only[a] | 1.25 | [0.98 ,1.59] | 0.07 | 4138 | 0.84 | [0.32 ,2.18] | 0.714 | 0.74 | [0.29 ,1.93] | 0.543 | 735 |

[a]Compared to spouses with non-migrating spouses

Note: Prevalence ratios (PR) presented with 95% confidence intervals (CI), spouse regressions adjusted for demographics except for marital status.

**Table S5 Crude and adjusted prevalence ratios for HIV and viremia regressions for relationship between migrant and non-migrant stratified by fishing and inland community.**

| | HIV prevalence | | | | HIV viremia (>1000 copies/mL) | | | |
|---|---|---|---|---|---|---|---|---|
| | Fishing | | Inland | | Fishing | | Inland | |
| | Crude PR [95% CI] | Adjusted PR [95% CI] | Crude PR [95% CI] | Adjusted PR [95% CI] | Crude PR [95% CI] | Adjusted PR [95% CI] | Crude PR [95% CI] | Adjusted PR [95% CI] |
| **Men** | | | | | | | | |
| Spouse with migrant spouse[a] | 0.99 [0.83, 1.18] | 0.86 [0.71, 1.04] | 0.95 [0.71, 1.27] | 0.99 [0.71, 1.39] | 1.52 [0.99, 2.32] | 1.46 [0.87, 2.46] | 1.79 [1.10, 2.92] | 1.23 [0.71, 2.14] |
| Parent with migrant child[b] | 1.16 [0.79, 1.71] | 0.73 [0.48, 1.10] | 1.91 [1.43, 2.54] | 0.95 [0.70, 1.30] | 0.62 [0.16, 2.38] | 1.02 [0.27, 3.91] | 0.63 [0.31, 1.28] | 1.00 [0.47, 2.13] |
| Sibling with migrant sibling[c] | 1.85 [0.59, 5.80] | 0.92 [0.37, 2.29] | 0.85 [0.40, 1.82] | 0.51 [0.26, 1.02] | . . | . . | . . | . . |
| **Women** | | | | | | | | |
| Spouse with migrant spouse[a] | 1.33 [1.08, 1.63] | 1.20 [0.97, 1.47] | 2.17 [1.66, 2.85] | 1.79 [1.37, 2.33] | 0.74 [0.28, 2.01] | 0.70 [0.25, 1.93] | 1.04 [0.43, 2.48] | 1.15 [0.47, 2.82] |
| Parent with migrant child[b] | 1.35 [1.10, 1.65] | 0.97 [0.79, 1.20] | 1.57 [1.33, 1.84] | 0.96 [0.81, 1.13] | 0.16 [0.02, 1.15] | 0.27 [0.04, 2.00] | 0.22 [0.08, 0.59] | 0.34 [0.12, 1.01] |
| Sibling with migrant sibling[c] | 2.10 [1.12, 3.92] | 1.53 [0.79, 2.96] | 1.30 [0.77, 2.21] | 0.84 [0.51, 1.39] | . . | . . | . . | . . |

[a]Compared to spouses with non-migrating spouses
[b]Compared to parents with non-migrating children
[c]Compared to siblings with non-migrating siblings
Note: Prevalence ratios (PR) presented with 95% confidence intervals [95% CI], spouse regressions adjusted for except for marital status; parent regressions adjusted for demographics including marital status. Sibling regressions excluded due to limited sample size.

**Table S6 Crude and adjusted prevalence ratios for HIV and HIV viremia by relationship between migrant and non-migrant stratified by direction of migration (into or out).**

| | Exposed n/N | Reference n/N | Crude PR [95%CI] | p | Adjusted PR [95%CI] | p |
|---|---|---|---|---|---|---|
| **HIV prevalence** | | | | | | |
| **Men** | | | | | | |
| Spouse with in-migrant spouse[a] | 91/452 | 482/2754 | 1.15 [0.94, 1.41] | 0.17 | 0.94 [0.76, 1.15] | 0.53 |
| Spouse with out-migrant spouse[a] | 95/372 | 482/2754 | 1.46 [1.20, 1.77] | **<0.01** | 0.89 [0.53, 0.76] | 1.15 |
| Parent with in-migrant child[b] | 24/76 | 487/3972 | 2.58 [1.83, 3.62] | **<0.01** | 1.16 [0.79, 1.70] | 0.45 |
| Parent with out-migrant child[b] | 45/278 | 487/3972 | 1.32 [1.00, 1.75] | **0.05** | 0.83 [0.61, 1.11] | 0.21 |
| Sibling with in-migrant sibling[c] | 5/81 | 29/1067 | 2.27 [0.90, 5.71] | 0.08 | 0.69 [0.30, 1.61] | 0.39 |
| Sibling with out-migrant sibling[c] | 9/422 | 29/1067 | 0.78 [0.37, 1.64] | 0.52 | 0.54 [0.27, 1.06] | 0.07 |
| **Women** | | | | | | |
| Spouse with in-migrant spouse[a] | 44/138 | 690/3991 | 1.84 [1.43, 2.38] | **<0.01** | 1.22 [0.94, 1.57] | 0.13 |
| Spouse with out-migrant spouse[a] | 55/147 | 690/3991 | 2.16 [1.74, 2.70] | **<0.01** | 1.57 [1.27, 1.95] | **<0.01** |
| Parent with in-migrant child[b] | 59/172 | 1030/5690 | 1.89 [1.53, 2.35] | **<0.01** | 1.06 [0.85, 1.32] | 0.61 |
| Parent with out-migrant child[b] | 139/605 | 1030/5690 | 1.27 [1.09, 1.48] | **<0.01** | 0.93 [0.79, 1.08] | 0.34 |
| Sibling with in-migrant sibling[c] | 12/76 | 53/935 | 2.79 [1.56, 4.98] | **<0.01** | 0.84 [0.50, 1.41] | 0.51 |
| Sibling with out-migrant sibling[c] | 22/316 | 53/935 | 1.23 [0.76, 1.99] | 0.40 | 0.99 [0.62, 1.59] | 0.97 |
| **HIV viremia** | | | | **-** | | |
| **Men** | | | | | | |
| Spouse with in-migrant spouse[a] | 26/89 | 86/480 | 1.63 [1.12, 2.38] | **0.01** | 1.32 [0.87, 2.00] | 0.19 |
| Spouse with out-migrant spouse[a] | 26/94 | 86/480 | 1.54 [1.06, 2.26] | **0.02** | 1.46 [0.91, 2.35] | 0.12 |
| Parent with in-migrant child[b] | 5/23 | 105/484 | 1.00 [0.45, 2.22] | 1.00 | 1.50 [0.73, 3.11] | 0.27 |
| Parent with out-migrant child[b] | 6/44 | 105/484 | 0.63 [0.29, 1.35] | 0.23 | 1.00 [0.43, 2.31] | 0.99 |
| Sibling with in-migrant sibling[c] | 1/5 | 12/29 | . | . | . | . |
| Sibling with out-migrant sibling[c] | 3/9 | 12/29 | . | . | . | . |
| **Women** | | | | | | |
| Spouse with in-migrant spouse[a] | 2/43 | 75/690 | 0.43 [0.11, 1.69] | 0.22 | 0.48 [0.12, 1.88] | 0.29 |
| Spouse with out-migrant spouse[a] | 7/55 | 75/690 | 1.17 [0.57, 2.42] | 0.67 | 1.15 [0.54, 2.47] | 0.71 |
| Parent with in-migrant child[b] | 1/58 | 130/1027 | 0.14 [0.02, 0.96] | 0.05 | 0.19 [0.03, 1.32] | 0.09 |
| Parent with out-migrant child[b] | 4/139 | 130/1027 | 0.23 [0.09, 0.61] | **<0.01** | 0.40 [0.14, 1.16] | 0.09 |
| Sibling with in-migrant sibling[c] | 1/12 | 10/53 | . | . | . | . |
| Sibling with out-migrant sibling[c] | 6/22 | 10/53 | . | . | . | . |

[a]Compared to spouses with non-migrating spouses; [b]Compared to parents with non-migrating children; [c]Compared to siblings with non-migrating siblings

Note: Prevalence ratios (PR) presented with 95% confidence intervals (CI), spouse regressions adjusted for demographics except for marital status; parent and sibling regressions adjusted for demographics including marital status. Sibling regressions for HIV viremia were excluded due to limited sample size. Bolded p-values <0.05
